# Real-Time Semi-Automated and Automated Voxel Placement for Repeated Acquisition Magnetic Resonance Spectroscopy

**DOI:** 10.1101/2021.09.23.21264046

**Authors:** James H. Bishop, Andrew Geoly, Naushaba Khan, Claudia Tischler, Ruben Krueger, Heer Amin, Laima Baltusis, Hua Wu, David Spiegel, Nolan Williams, Matthew D. Sacchet

## Abstract

Magnetic resonance spectroscopy (MRS) is heavily dependent on the investigative team to prescribe, or demarcate, the desired tissue volume-of-interest. Manual prescription, the current standard in the field, requires expertise in neuroanatomy to ensure spatial consistency within and across subjects. Spatial precision of MRS voxel placement thus presents challenges for cross-sectional studies, and even more so for repeated-measure and multi-acquisition designs. Furthermore, voxel prescriptions based-solely on anatomical landmarks may not be ideal in regions with substantial functional and cytoarchitectural variability or to examine individualized/targeted interventions. Here we propose and validate robust and real-time methods to automate MRS voxel placement using functionally defined coordinates within the left dorsolateral prefrontal cortex in clinical cohorts of chronic pain and depression. We hypothesized that increased automation would produce more consistent voxel placement across repeated acquisitions particularly in reference to standard manual prescription. Data were collected and analyzed using two independent prospective transcranial magnetic stimulation studies: 1) a single-day multi-session sandwich design and 2) a longitudinal design. Participants with fibromyalgia syndrome (N=50) and major depressive disorder (N=35) underwent MRI as part of ongoing clinical studies. MEGA-PRESS and Optimized-PRESS MRS acquisitions were acquired at 3-tesla. Evaluation of the reproducibility of spatial location and tissue segmentation was assessed for: 1) manual, 2) semi-automated, and 3) automated voxel prescription approaches. Variability of grey and white matter voxel tissue composition was reduced using automated placement protocols as confirmed by common MRS software processing pipelines (Gannet; SPM-based segmentation) and via Freesurfer-based segmentation. Spatially, post-to pre-voxel center-of-gravity distance was reduced and voxel overlap increased significantly across datasets using automated compared to manual procedures. These results demonstrate the within subject reliability and reproducibility of a method for reducing variability introduced by spatial inconsistencies during MRS acquisitions. The proposed method is a meaningful advance toward improved consistency of MRS data in neuroscience and can be leveraged for multi-session and longitudinal studies that target precisely defined regions-of-interest via a coordinate-based approach.

## INTRODUCTION

Magnetic resonance spectroscopy (MRS) is a non-invasive brain imaging approach capable of quantifying diverse metabolic and biochemical processes including specific neurotransmitters (1,2). For many other magnetic resonance imaging (MRI) approaches it is often standard procedure to collect data from the entire brain and then conduct *post hoc* analyses on defined regions-of-interest. Alternatively, MRS generally requires the user to prospectively delineate the tissue volumes-of-interest (VOI). This difference in MRS acquisition necessitates new methods for reliable and reproducible data collection that are unique from those for other imaging approaches (e.g., fMRI). Designing studies without precisely placed voxels limits the utility of MRS for use in evaluating complex disorders and interventions. As a related example, neuromodulation approaches such as transcranial magnetic stimulation (TMS) have been historically guided by skull-based measurements. Prefrontal structures targeted for clinical applications demonstrate substantial variability across individuals (3). Even with the capability of modulating brain tissue with centimeter-resolution (4), evidence suggests that individualized optimization of treatment location using functional brain imaging constrained within a designated anatomical region provides more consistent therapeutic outcomes (5,6). Similarly, investigating chemical alterations using MRS defined by standard anatomical landmarks across individuals may limit the utility and interpretability of such findings.

Precision of MRS VOIs is generally reliant on the clinical or investigative team and their neuroanatomical expertise. Furthermore, consistent voxel prescription is dependent on inter-subject anatomical variability. These factors may be compounded as the size of the VOI decreases, and with it the related biochemical measurements of interest (7). Standard MRS VOI placement approaches are especially problematic for multi-center, cross-sectional, and repeated measures trials that involve data collection by multiple users over time. Indeed, long-term projects often have turnover in research staff. Taken together, these factors contribute to increased variability and decreased consistency of voxel placement and thus weaken the validity of MRS measurements.

Inter-individual anatomical variability is a challenge for voxel prescription. Even if spatial consistency is achieved manually by the MRI system operator using only grossly visible landmarks, this does not ensure that the user is measuring the *functionally* analogous region across participants. Many of the current approaches for VOI placement ignore the functional and/or cytoarchitectural heterogeneity across brain structures because they are not visualizable. For context, prefrontal structures such as the dorsolateral prefrontal cortex (DLPFC) are highly variable across participants and have undergone iterations of refinement since the original Brodmann parcellations (8-11). This further confounds the utility of MRS to investigate complex disorders where the neural circuitry involved may not demonstrate grossly observable pathology. While integration of real-time functional MRI tasks for guiding voxel prescription has been developed, these techniques require robust and validated tasks that must be implemented in a short timeframe. For example, the use of functional localizers and task-based methods such as finger tapping, the n-back task, or visual stimuli have been use to guide voxel placement in the motor, prefrontal, and visual cortices respectively (12-14). These methods are promising, however, an expansion of the approach to enable coordinate-based prescription across participants is warranted for placement in regions that lack definitive task-based activation paradigms or to ensure consistent placement across repeated MRI visits/scans without needing to reacquire localizers. Coordinate based voxel prescription enables a wide array of methodological flexibility that may increase the reliability within participants and across studies.

Coordinate based anatomical voxel prescription methods have been developed to overcome the variability introduced by manual voxel placement, but the current approaches have yet to integrate placement with functional targets or in a within participant registration paradigm for repeated acquisition consistency. For example, several methods have been proposed that afford user independent automated VOI placement that utilize brain co-registration (alignment) methods. These rely on either affine (linear) or b-spline (non-linear) brain co-registration that align a subject’s anatomical scan to a standard brain atlas or template during the imaging session (15). The registration parameters are then used to quantitatively identify template-to-individual voxel placement coordinates which are then input by the user into the scanner acquisition software. Within automated approaches the choice of registration method has been shown to influence accuracy (16); however, this is a greater issue when registering standard brain templates or atlases to an individual subject’s T1-weighted (T1w) image. For template-to-subject registration, non-linear registration approaches outperform affine registration at the downfall of increased computational time and required computing power (17). Alternatively, affine approaches are faster and require less computational resources (which may be helpful in real-time data acquisition contexts), and the performance of these approaches are optimal for within subject coregistration, however, this requires repeated acquisition paradigms.

The use of MRS as a neuroscientific tool for the identification of neurochemical concentrations will benefit from methods that can be conducted quickly by study personnel, are reliably prescribed in an automated fashion, applied on an individual subject basis, and are reproducible across longitudinal, multi-acquisition, and repeated measurements. The methods used to determine VOIs, whether based on function, structure, or otherwise, are ultimately dependent on researcher or clinician preference and the scientific/clinical question. Here we provide a methodological framework for the automation of repeated-measure and longitudinal acquisition of MRS voxels in a heterogenous functional brain region, the dorsolateral prefrontal cortex (DLPFC). In this investigation, we examine two iterations of our approach which we term “semi-automated” and “automated” based on the amount of user input across repeated measure and longitudinal acquisitions as well as a manual MRS voxel prescription in two independent clinical datasets. We leverage fast b-spline (affine) registration of anatomical images and co-registration of an individual-based functional (fMRI) coordinate of interest to center the VOI in a clinically relevant portion of the left DLPFC (L-DLPFC). We hypothesized that the semi-automated and automated approaches would reduce spatial and tissue segmentation variability across repeated MRS acquisitions and voxels of different sizes within the L-DLPFC, as compared to a standard manual MRS voxel prescription approach. We implemented field-standard segmentation approaches, including those packaged within Freesurfer (18) and Statistical Parametric Mapping (SPM) (19), to evaluate tissue fraction across prescription pipelines. To quantify the consistency of voxel placements, we calculated Euclidean distance from the center-of-gravity coordinates across scans, in addition to determining the similarity of MRS voxels using an overlap coefficient across repeated MRS acquisitions.

## METHODS

### Participant Recruitment and Evaluation

Participants were recruited as part of two separate studies. Both studies were approved by the Stanford University Institutional Review Board (IRB) and participants provided informed consent. All procedures were in compliance with the Declaration of Helsinki. Prior to imaging, participants were evaluated for standard MRI contraindications, in-person, by study coordinators and a study physician and provided informed consent.

Study A was used to evaluate the voxel placement between independent MRI sessions spaced one hour apart. In brief, 50 participants with fibromyalgia syndrome, a chronic pain disorder, first underwent a baseline MRI (MRI #1) followed by two separate experimental days - each with two independent MRI scans (MRI #2-5) sandwiched between transcranial magnetic stimulation treatment (**Fig. 1**). Resting state functional scans were acquired at the baseline MRI (MRI #1), analyzed, and the identified L-DLPFC cluster coordinate was used during each of the subsequent MRIs (MRI #2-5) for automated voxel prescription. MRI sessions were approximately 1-hours and included a combination of structural, chemical, and functional acquisitions. GABA-edited MEGA-PRESS (MEshcher-Garwood Point RESolved Spectroscopy) and broad spectra Optimized-PRESS MRS sequences were collected pre- and post-TMS MRI sessions only. Imaging parameters are described in greater detail below. A total of 50 pre-/post-MEGA-PRESS and 50 pre-/post-Optimized-PRESS scans were collected culminating in 200 total acquisitions.

**Figure 1.**
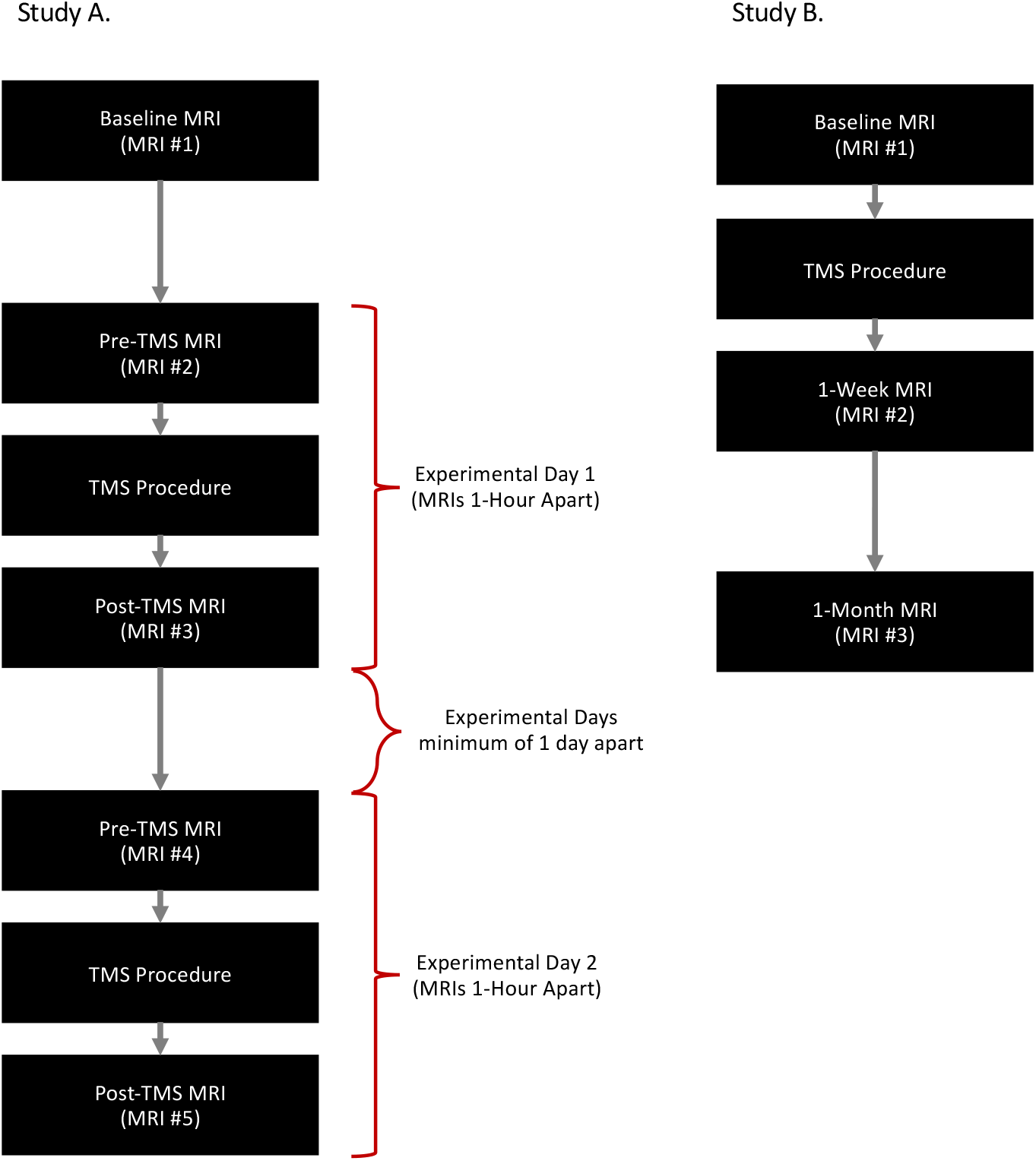
Experimental Design: Study A (left) timeline demonstrating a repeated-acquisition sandwich design where there were two different imaging days consisting of two independent MRI scans sandwiched between a TMS intervention. Study B (right) demonstrates a longitudinal imaging design with three independent scans at varying follow-up timeframes: baseline, one-week, and one-month MRI visits.

Study B was conducted in a treatment resistant major depressive disorder population undergoing resting state functional connectivity guided TMS therapy. The paradigm was used to evaluate within-subject voxel placements between three independent MRI sessions at the following timepoints: (1) baseline (MRI #1), (2) one-week (MRI #2), and one-month (MRI #3, **Fig. 1**). Resting state functional scans were collected and analyzed during the baseline MRI (MRI #1) and thus manual DLPFC voxel placement was instituted during this timepoint. The identified L-DLPFC resting state cluster coordinate was then used to guide the voxel placement during the one-week and one-month MRI evaluations (MRI #2-3). A combination of structural, chemical, and functional imaging was collected at all MRI sessions. MEGA-PRESS MRS for the 38 participants was acquired at each of the three timepoints detailed above and included 114 total MEGA-PRESS acquisitions. Imaging parameters are described in greater detail below.

### MRI Data Acquisition

MRI data were obtained using a research dedicated 3.0T General Electric Discovery MR750 instrument with a Nova Medical 32-channel head coil. Acquisition parameters were identical across both studies (i.e., studies A & B) and included a combination of: structural, chemical, and functional acquisitions. Whole-brain structural imaging consisted of a 0.9 mm^3^ three-dimensional T1w Magnetization Prepared - Rapid Gradient Echo (MPRAGE) sequence. Whole-brain high resolution fMRI (resting state) was collected using a simultaneous multi-slice EPI sequence with the following parameters: echo time (TE) = 30 ms, repetition time (TR) = 2 s, flip angle = 77°, slice thickness = 1.8 mm, and FOV = 230 mm. Broad spectra and GABA+ MRS data were collected using Optimized-PRESS (20-22) and MEGA-PRESS (23) sequences respectively within the left DLPFC (L-DLPFC). MEGA-PRESS sequence parameters included: voxel size = 20×20×20 mm^3^ (8 mL), TE = 68 ms; TR = 2 s; editing pulses applied at 1.9 ppm (ON) and 7.46 ppm (OFF) for a total acquisition time of ∼10 min. Optimized-PRESS sequence parameters included: voxel size = 14×14×14 mm^3^ (2.744 mL), TE = 35 ms; TR = 2 s for a total acquisition time of ∼3 min. MRS voxels were placed according to several strategies described below.

### Identification of rs-fMRI cluster for guided voxel prescription

While both structural and functional targets are compatible with our automated MRS voxel placement procedure, here we investigated the reliability and utility of MRS targets that were defined functionally given previous methods have validated the utility of structurally defined approaches using atlas-based coordinates.

In both studies, resting state functional connectivity clusters were identified to guide clinical TMS therapy for either chronic pain (Study A) or depression (Study B) providing a relevant paradigm for future application of this automated voxel prescription technique. In Study A, a voxelwise analysis of the rs-fMRI scan was analyzed to determine the subregion of the L-DLPFC (Brodmann Area 9 + 46) that exhibited the greatest correlation with the dorsal anterior cingulate (dACC) (24). In Study B, a voxelwise analysis of the rs-fMRI scan was analyzed to determine the subregion of the L-DLPFC (Brodmann Area 46) exhibiting the greatest anti-correlation to the subgenual cingulate (sgCC) (5).

### Voxel Prescription Procedures

In the two studies we implemented three voxel prescription approaches: 1) manual, 2) semi-automated, and 3) automated. Each of the study specific protocols are outlined in detail below and typically can be completed concurrently during other desired acquisitions (i.e., structural or additional spectroscopic acquisitions, etc.) in several minutes or less:

In Study A, semi-automated and automated approaches were used. The baseline resting state fMRI scan (from the MRI #1; **Fig. 2**) was first analyzed as described above. On the subsequent independent pre- and post-TMS MRI sessions, L-DLPFC Optimized-PRESS and MEGA-PRESS voxel prescription was performed using two different iterations of our voxel prescription procedure termed semi-automated and an automated based on amount of required user input and fine tuning. For both prescription approaches, the center of gravity coordinate (mm) of the identified L-DLPFC functional cluster was extracted using the FSL Software (Version 6.0) via the fslstats function. In the semi-automated voxel placement approach, participants then underwent a pre-TMS MRI. During this imaging session, a T1w image was collected prior to either MEGA-PRESS, Optimized-PRESS, or both. Immediately following completion of the T1w image acquisition, a custom script was used to pull the MRI data directly from the imaging server. The T1w image was then reconstructed into Neuroimaging Informatics Technology Initiative (NIFTI) format. Next, a second in-house MATrix LABoratory (MATLAB; R2015a, The Mathworks, Inc.) script utilizing Statistical Parametric Mapping software version 12 functions (SPM12; Wellcome Trust Centre for Neuroimaging). The baseline (MRI #1) T1w and pre-TMS (MRI #2) T1w images were co-registered while the participant underwent additional study specific MRI acquisitions. Co-registration was achieved using the spm_coreg.m function. Affine transformation matrices were defined using an optimized normalized mutual information approach (25-27). Resulting affine transformation matrices were then used to convert the fMRI-based L-DLPFC center-of-gravity coordinate from baseline (MRI #1) to pre-TMS MRI space.

**Figure 2.**
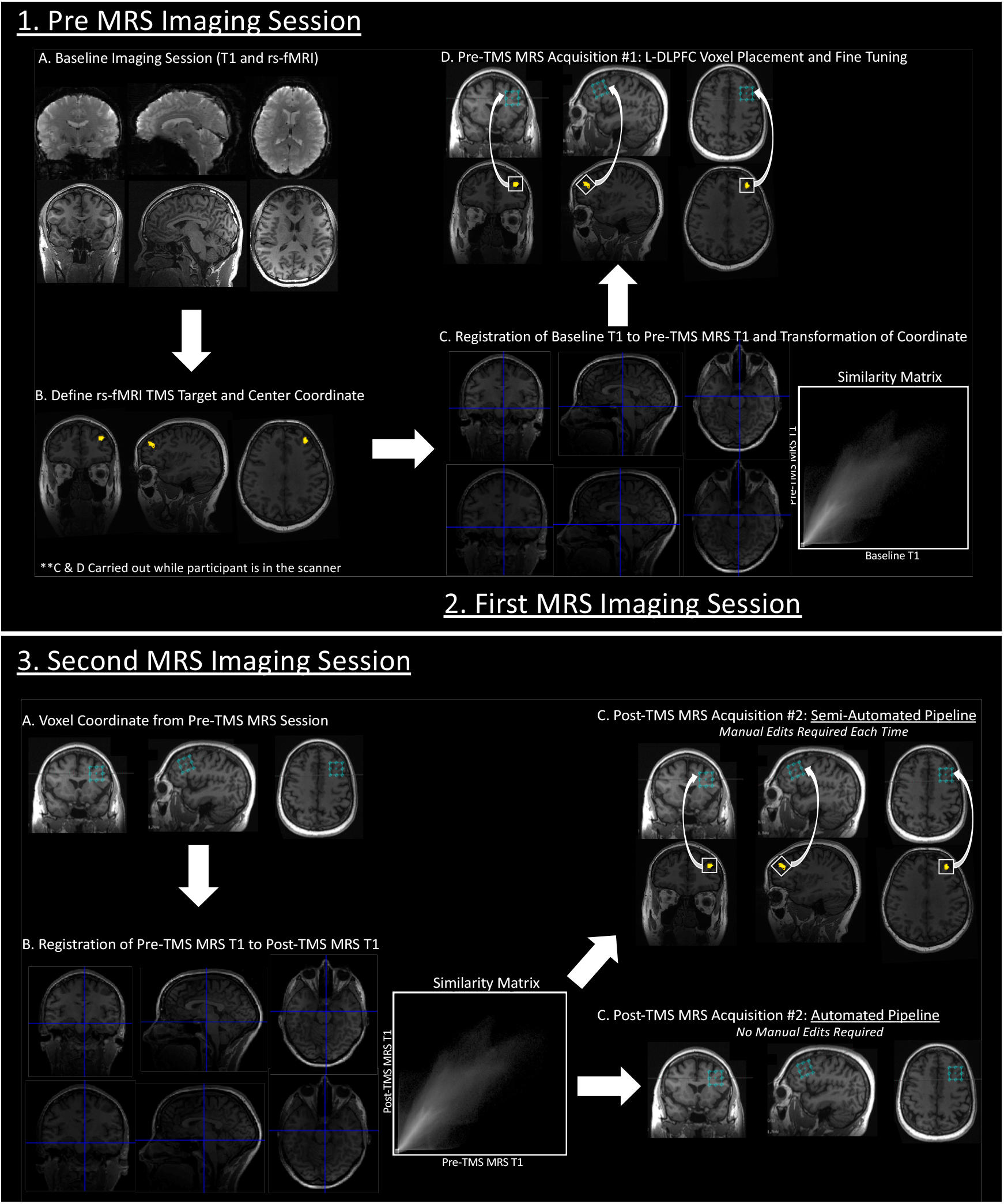
VOI Prescription Schematic: Schematic of semi-automated and automated voxel prescription procedure. Step 1 (upper section of image) – Pre-MRS Imaging Session: An MRI session consisting of T1w and resting state acquisitions (Study A) and also a manually prescribed L-DLPFC MEGA-PRESS scan for Study B (A). Resting state images were preprocessed and analyzed to define a functional ROI within the L-DLPFC (B). The center-of-gravity coordinate for the functional cluster was then determined using FSL tools (fslstats). 2 – First MRS Imaging Session: During the second imaging session, a T1w was first acquired. Upon completion the scan, the T1w image was pulled directly from the MR system server, reconstructed, and co-registered to the pre-MRS T1w image (C). MRS voxel rotation aligned with the slope of the skull in the sagittal plane and the co-registered center-of-gravity coordinate in the current subject space was input into the scanner. Voxel location adjustments were made to ensure the entirety of the voxel was within brain tissue and did not encompass meninges or skull (D). Following minor adjustments, the final center-of-gravity voxel coordinates were documented for use in the subsequent independent MRS sessions. Notably this entire procedure requires several minutes (<3 in our experience). 3 – Second MRS Imaging Session: For the semi-automated voxel prescription pipeline the identical procedure described above was repeated (not shown). However, for the automated voxel prescription pipeline, the T1w image was again acquired first, directly pulled from the MR system server, and reconstructed. This time the current T1w image was co-registered to the first MRS T1w image along with the documented center-of-gravity coordinate defined after adjustments were made. Voxel rotation was aligned with the slope of the skull in the sagittal plane and the computed coordinate was input into the scanner software. If done correctly no further manual modification to the voxel location was required.

Pre-TMS MRI voxel prescription for both Optimized-PRESS and MEGA-PRESS was then setup in two steps. First, voxel rotations were aligned to the skull geometry in the sagittal plane using a shim acquisition. Once the appropriate rotation was achieved and copied to either the MEGA-PRESS or Optimized-PRESS sequence, the voxel size was specified (MEGA-PRESS = 20×20×20 mm^3^ and Optimized-PRESS 14×14×14 mm^3^) and the co-registration-defined coordinate (described above) was then input into the scanner console interface. This resulted in the placement of the center of the MRS voxel at the center of the desired functional ROI (**Fig. 2**). The term semi-automated pipeline was used due to the superficial nature of the cortical targets which required manually translating the voxel to ensure only brain tissue was encompassed within the bounding box (i.e., not skull and other non-brain tissue that can influence the MRS measurement). For the semi-automated pipeline group, this process, with the slight manual translation, was then repeated for all subsequent MRI sessions (**Fig. 2D top**).

To further remove the required translation step from all but the first MRS acquisition, an automated procedure was developed. This automated method first implements the semi-automated approach as described above for registration from MRI #1 to MRI #2, followed by an increasingly automated procedure that obviates the need to translate the MRS voxel to avoid non-brain tissue. This is achieved by directly co-registering the translated MRS coordinate from MRI #2 for all subsequent acquisitions. The output of this procedure is thus a new co-registered coordinate that does not require additional manual adjustment for any subsequent repeated acquisition (**Fig. 2**).

In Study B, MRS (MEGA-PRESS only) scans were acquired at baseline (MRI #1), after one week, and again after one month (**Fig. 2**) using manual or semi-automated voxel prescription approaches. Voxel prescription at baseline was conducted manually according to neuroanatomy without the use of any semi- or automated voxel placement procedures. In brief, the voxel was aligned to the angle of the skull in the sagittal plane. Co-registration of the Brodmann Area 46 mask in standard space to the subjects T1-weighted image was first conducted and then the voxel was manually transcribed to center of the mask by visualizing participant specific anatomical landmarks. The 1-week and 1-month voxels were prescribed using the semi-automated approach described above. (**Fig. 2**).

### Voxel Composition

GM and WM voxel segmentation fractions were evaluated across prescription protocols to determine the consistency and reliability of semi-automated and automated approaches. Both Freesurfer and the SPM segmentation output which is standard in the Gannet MRS processing package were assessed.

#### Freesurfer Analysis

T1w images were segmented using Freesurfer software (Version 6; (18)). Next, a custom MATLAB script was utilized to extract the 3-dimensional (3D) voxel mask from the raw MRS data (i.e., GE p-file). The scanner-reconstructed T1w image was then reoriented to standard space (fslreorient2std; FSL software toolbox) and the 3D voxel mask and script-generated T1w image geometries were standardized using the flscpgeom command. AFNI (28) 3dcalc was then used to compute the WM, GM, and cerebrospinal fluid (CSF) segmentation percentages of the 3D voxel from the Freesurfer generated segmentation (aseg.mgz) file.

#### SPM Analysis

The Gannet Software toolbox (29) is a freely available software suite that is commonly used to process and analyze MEGA-PRESS data. Gannet processing includes the option to implement batch tissue segmentation using SPM12 Software (19). GM, WM, and CSF tissue fractions were extracted from the MEGA-PRESS p-files using SPM via the GannetSegment function in the Gannet software toolbox (Version 3.0). Subsequent statistical analyses were carried out on GM and WM tissue fractions.

Although it is not within the scope of this manuscript to directly compare tissue segmentation approaches (i.e., Freesurfer vs. SPM), tissue fraction differences within the MRS voxel are known to influence metabolic concentration (30,31). For this reason, both Freesurfer and SPM segmentations were generated and compared.

### Spatial Consistency of Voxel Placement

#### Euclidean Distance Analyses

Euclidean distance from the center-of-gravity voxel coordinates across MRI timepoints were calculated to evaluate the stability of each of the prescription protocols (i.e., Study A MEGA-PRESS semi-automated vs. automated; Study A Optimized-PRESS semi-automated vs. automated; Study B MEGA-PRESS manual vs. semi-automated). Distance analyses were conducted in subject (T1w) space using FSL Software. For both Study A and Study B protocols, subjects’ T1w scans for all MRI timepoints (MRI #2 and MRI #3) were linearly registered (flirt) to the baseline MRI (MRI #1) for both Study A and Study B. Next, the MRS voxels for each subsequent timepoint were co-registered to the baseline MRI scan. Three-dimensional center-of-gravity coordinates (mm-space) were extracted for each voxel and Euclidian distance was calculated.

#### Dice Similarity Coefficient

To examine the spatial overlap of the MRS voxels, the Sørensen–Dice Similarity Coefficient (DSC) was implemented (32,33). This is also referred to as the dice overlap coefficient. Each subjects T1w scans were co-registered to their baseline T1w scan using FSL LInear Registration Tools (FLIRT) with no resampling of the MRS voxels. DSC was then calculated in RStudio (*RStudio Team (2020). RStudio: Integrated Development for R. RStudio, PBC, Boston, MA URL* http://www.rstudio.com/) with the FSL wrapper functions (https://rdrr.io/cran/fslr/man/fsl_dice.html). DSC was computed across all pipelines using binarized pre- and post-MRS voxel masks which are represented in the equation below as A and B respectively. DSC outputs range from 0, representing no overlap, to 1 which represents complete overlap. Greater overlap indicates greater consistency across the multi-session data.

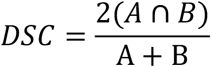

### Statistical Analyses

All statistical analyses were carried out in RStudio (Version 2018 1.2.1335). To determine the appropriate downstream statistical test to investigate differences in variability (SD) and means (M), both homogeneity of variance and normality were first assessed for segmentations (GM and WM), Euclidean distance, and overlap coefficient variables. Normality (distribution) of each variable was evaluated using the Shapiro-Wilk normality test (34). Homogeneity of variances were assessed between groups using either a standard *F* test if the variable(s) were normally distributed or alternatively with the non-parametric Fligner-Killeen Test (35) if there was a significant deviation from normality (*p* < 0.5). Between group mean differences were determined using unpaired two-sample *t*-tests in Specifically, Welch’s *t*-tests were implemented when homogeneity of variance tests were significant (p < 0.5).

## RESULTS

### Voxel Composition

GM and WM composition of the L-DLPFC VOIs was assessed by first calculating the difference in longitudinal tissue fraction (Post-Pre) across prescription pipelines using standard segmentation approaches including both Freesurfer and SPM (**Table 1 & Fig. 3**). In Study A (MEGA-PRESS: *N* = 50; semi-automated = 26, automated = 24; Optimized-PRESS: *N* = 50; semi-automated = 19, automated = 31), individuals underwent longitudinal MRI scanning approximately one-hour apart and semi-automated voxel prescription was compared to automated voxel prescription. Three participants in each of Study A and Study B were excluded from the analyses due to poor data quality and/or registration related issues (e.g., scanner related shim error). No significant between-pipeline differences in GM or WM voxel composition were identified for either MEGA-PRESS (voxel size = 20mm^3^) or Optimized-PRESS acquisitions (voxel size = 14mm^3^) across segmentation methods. A significant between-pipeline difference in variance was observed in the MEGA-PRESS acquisition for both Freesurfer and SPM segmentations (**Table 1**), demonstrating a reduction in variability in voxel composition of both GM and WM with the automated prescription approach. Alternatively, tissue composition of the smaller Optimized-PRESS acquisition did not yield significant between-pipeline differences in either mean or variance.

**Table 1.**
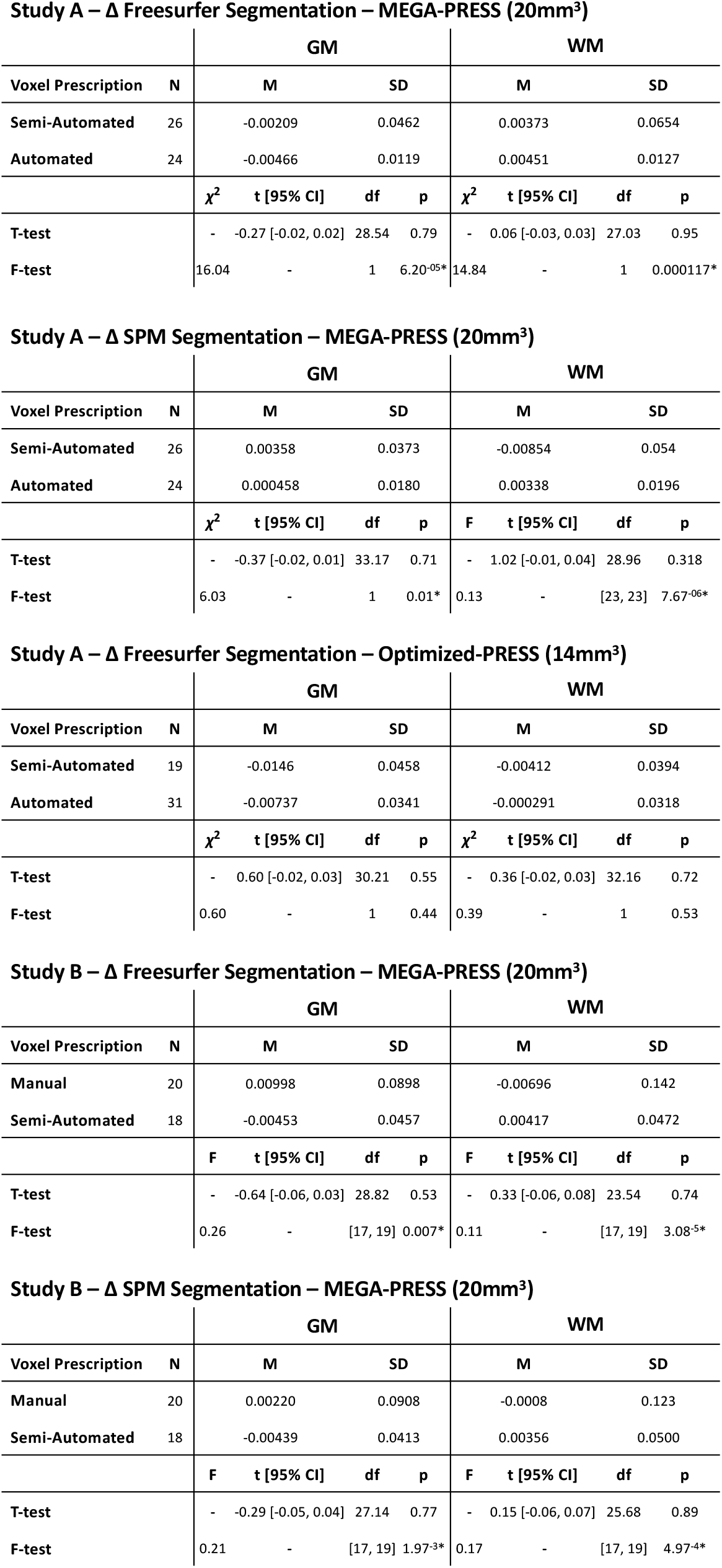
Voxel Composition Across Multi-Acquisition and Longitudinal Studies: Tissue fraction statistics for each project across voxel prescription approaches, MRS acquisitions of different sizes, and segmentation methods. Statistically significant differences are indicated by (*).

**Study A – Δ Freesurfer Segmentation – MEGA-PRESS (20mm<sup>3</sup>)**
|  |  | GM |  |  |  | WM |  |  |  |
| --- | --- | --- | --- | --- | --- | --- | --- | --- | --- |
| Voxel Prescription | N | M | SD |  |  | M | SD |  |  |
| Semi-Automated | 26 | -0.00209 | 0.0462 |  |  | 0.00373 | 0.0654 |  |  |
| Automated | 24 | -0.00466 | 0.0119 |  |  | 0.00451 | 0.0127 |  |  |
| | | $\chi^2$ | t [95% CI] | df | p | $\chi^2$ | t [95% CI] | df | p |
| T-test |  | - | -0.27 [-0.02, 0.02] | 28.54 | 0.79 | - | 0.06 [-0.03, 0.03] | 27.03 | 0.95 |
| F-test |  | 16.04 | - | 1 | 6.20<sup>-05*</sup> | 14.84 | - | 1 | 0.000117* |

**Study A – Δ SPM Segmentation – MEGA-PRESS (20mm<sup>3</sup>)**
|  |  | GM |  |  |  | WM |  |  |  |
| --- | --- | --- | --- | --- | --- | --- | --- | --- | --- |
| Voxel Prescription | N | M | SD |  |  | M | SD |  |  |
| Semi-Automated | 26 | 0.00358 | 0.0373 |  |  | -0.00854 | 0.054 |  |  |
| Automated | 24 | 0.000458 | 0.0180 |  |  | 0.00338 | 0.0196 |  |  |
| | | $\chi^2$ | t [95% CI] | df | p | F | t [95% CI] | df | p |
| T-test |  | - | -0.37 [-0.02, 0.01] | 33.17 | 0.71 | - | 1.02 [-0.01, 0.04] | 28.96 | 0.318 |
| F-test |  | 6.03 | - | 1 | 0.01* | 0.13 | - | [23, 23] | 7.67<sup>-06*</sup> |

**Study A – Δ Freesurfer Segmentation – Optimized-PRESS (14mm<sup>3</sup>)**
|  |  | GM |  |  |  | WM |  |  |  |
| --- | --- | --- | --- | --- | --- | --- | --- | --- | --- |
| Voxel Prescription | N | M | SD |  |  | M | SD |  |  |
| Semi-Automated | 19 | -0.0146 | 0.0458 |  |  | -0.00412 | 0.0394 |  |  |
| Automated | 31 | -0.00737 | 0.0341 |  |  | -0.000291 | 0.0318 |  |  |
| | | $\chi^2$ | t [95% CI] | df | p | $\chi^2$ | t [95% CI] | df | p |
| T-test |  | - | 0.60 [-0.02, 0.03] | 30.21 | 0.55 | - | 0.36 [-0.02, 0.03] | 32.16 | 0.72 |
| F-test |  | 0.60 | - | 1 | 0.44 | 0.39 | - | 1 | 0.53 |

**Study B – Δ Freesurfer Segmentation – MEGA-PRESS (20mm<sup>3</sup>)**
|  |  | GM |  |  |  | WM |  |  |  |
| --- | --- | --- | --- | --- | --- | --- | --- | --- | --- |
| Voxel Prescription | N | M | SD |  |  | M | SD |  |  |
| Manual | 20 | 0.00998 | 0.0898 |  |  | -0.00696 | 0.142 |  |  |
| Semi-Automated | 18 | -0.00453 | 0.0457 |  |  | 0.00417 | 0.0472 |  |  |
|  |  | F | t [95% CI] | df | p | F | t [95% CI] | df | p |
| T-test |  | - | -0.64 [-0.06, 0.03] | 28.82 | 0.53 | - | 0.33 [-0.06, 0.08] | 23.54 | 0.74 |
| F-test |  | 0.26 | - | [17, 19] | 0.007* | 0.11 | - | [17, 19] | 3.08<sup>-5*</sup> |

**Table 1. Voxel Composition Across Multi-Acquisition and Longitudinal Studies:**
|  |  | GM |  |  |  | WM |  |  |  |
| --- | --- | --- | --- | --- | --- | --- | --- | --- | --- |
| Voxel Prescription | N | M | SD |  |  | M | SD |  |  |
| Manual | 20 | 0.00220 | 0.0908 |  |  | -0.0008 | 0.123 |  |  |
| Semi-Automated | 18 | -0.00439 | 0.0413 |  |  | 0.00356 | 0.0500 |  |  |
|  |  | F | t [95% CI] | df | p | F | t [95% CI] | df | p |
| T-test |  | - | -0.29 [-0.05, 0.04] | 27.14 | 0.77 | - | 0.15 [-0.06, 0.07] | 25.68 | 0.89 |
| F-test |  | 0.21 | - | [17, 19] | 1.97<sup>-3*</sup> | 0.17 | - | [17, 19] | 4.97<sup>-4*</sup> |

**Figure 3.**
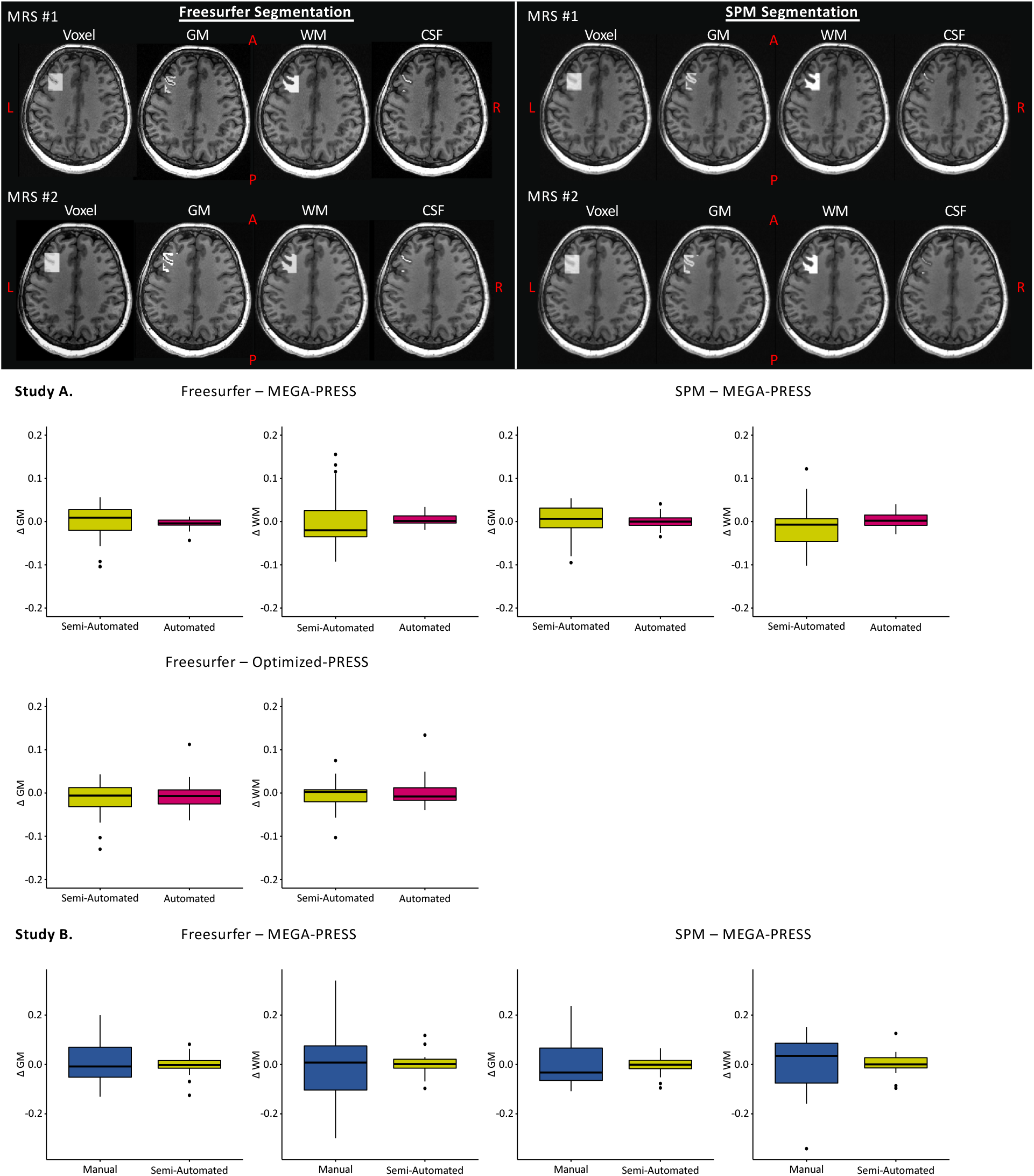
Multi-Acquisition / Longitudinal L-DLPFC Voxel Tissue Composition: Tissue fractions were determined by calculating the differences between Freesurfer and SPM segmentations across repeated independent MEGA-PRESS and Optimized-PRESS acquisitions of difference voxel sizes. In Study A, tissue delta was defined as Post MRI – Pre MRI GM and WM tissue fractions for both semi-automated and automated prescription pipelines (Top). Study B included only MEGA-PRESS acquisitions, with identical scan parameters as Study A. GM and WM tissue fractions for manual and semi-automated placement were determined by computing the difference between the one-week MRI – baseline MRI and one-month MRI – one-week MRI respectively. With the exception of the Study A Optimized-PRESS measures, the more automated prescription procedures reduced variability of tissue fraction.

In Study B (N=38; manual=20, semi-automated=18), individuals underwent longitudinal MRI scanning at three independent timepoints: baseline, one-week, and one-month. Manual prescription occurred at baseline and semi-automated functional connectivity guided placement occurred at one-week and 1-month utilizing the same functional-connectivity derived coordinate. Similar to Study A, the mean GM and WM fractions were not significantly different between voxel placement pipelines for either segmentation approach (**Fig. 3**). Also consistent with Study A, a significant between-pipeline difference in variance was identified across voxel prescription pipelines and segmentation approaches (**Table 1 & Fig. 3**).

### Spatial Consistency: Euclidean Distance & DSC

Euclidean distance was calculated and compared across acquisitions to assess the movement of the center-of-gravity for the MRS voxel (**Fig. 5**). DSC was computed and compared to further characterize the extent of spatial overlap between voxel prescription pipelines (**Fig. 4**). Three participants in each of Study A and Study B were excluded from the analyses due to poor data quality and/or registration related issues (e.g., scanner related shim error). In Study A, the automated voxel prescription performed significantly better than semi-automated placement for both MEGA-PRESS and Optimized-PRESS acquisitions as indicated by a reduction in center-of-gravity Euclidean distance and increased spatial overlap across repeated scans. This was demonstrated by large effect sizes across statistical comparisons of both Euclidian distance and DSC: MEGA-PRESS Euclidean distance (Cohen’s d = -1.76, 95% CI = [-2.41, -1.10]), MEGA-PRESS DSC (Cohen’s d = 1.75, 95% CI = [1.09, 2.40]), Optimized-PRESS Euclidean distance (Cohen’s d = -1.59, 95% CI = [-2.04, -1.13]), and Optimized-PRESS DSC (Cohen’s d = 1.38, 95% CI = [0.94, 1.82]). Similarly, in Study B, semi-automated voxel prescription significantly outperformed manual placement in both center-of-gravity Euclidean distance and spatial overlap (DSC) of the voxels even with the increased duration between scans, that is: one-month vs. one-week. Effect sizes were also respectively large: MEGA-PRESS Euclidean distance (Cohen’s d = -2.25, 95% CI = [- 3.10, -1.38]) and MEGA-PRESS DSC (Cohen’s d = 2.42, 95% CI = [1.52, 3.29]).

**Figure 4.**
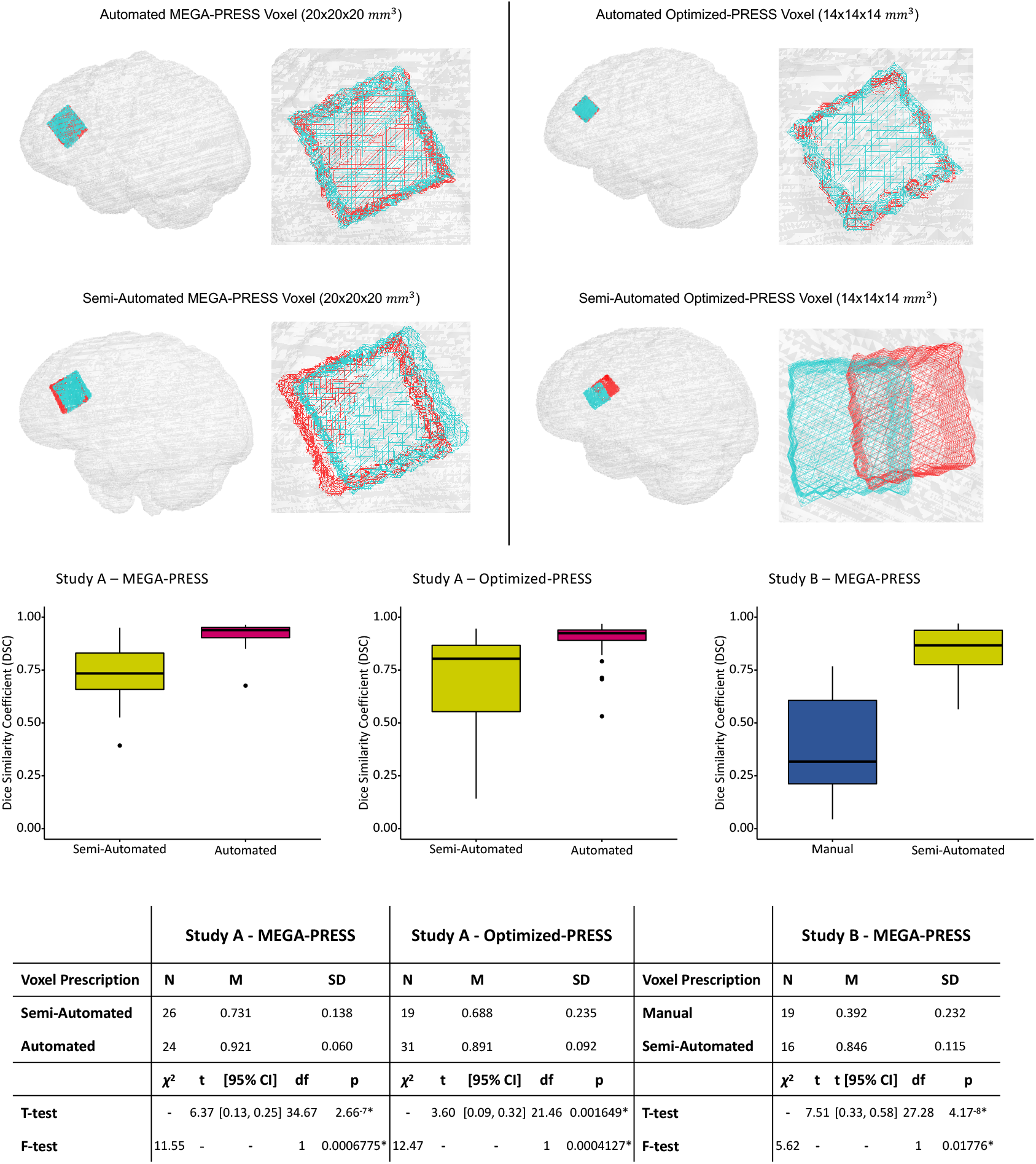
L-DLPFC VOI Spatial Overlap. Spatial congruency of the repeated MRS voxel prescriptions was also assessed by computing a dice similarity coefficient DSC). Analyses were completed in individual subject space (T1w image-space) without the need for resampling the MRS voxel. Outputs of the DSC range from 0 (no overlap) to 1 (complete overlap). The table illustrates that the more automated the voxel prescription procedure the more spatial overlap regardless of voxel size. This is consistent in Study A where independent scans were acquired an hour apart and the automated pipeline outperformed the semi-automated prescription procedure with regard to overlap of the repeated placement. In Study B, the semi-automated prescription procedure outperformed the manual placement procedure even with increased duration between scans in the automated data acquisition, that is: Manual prescription data were acquired one-week apart and semi-automated data were acquired 1-month apart.

**Figure 5.**
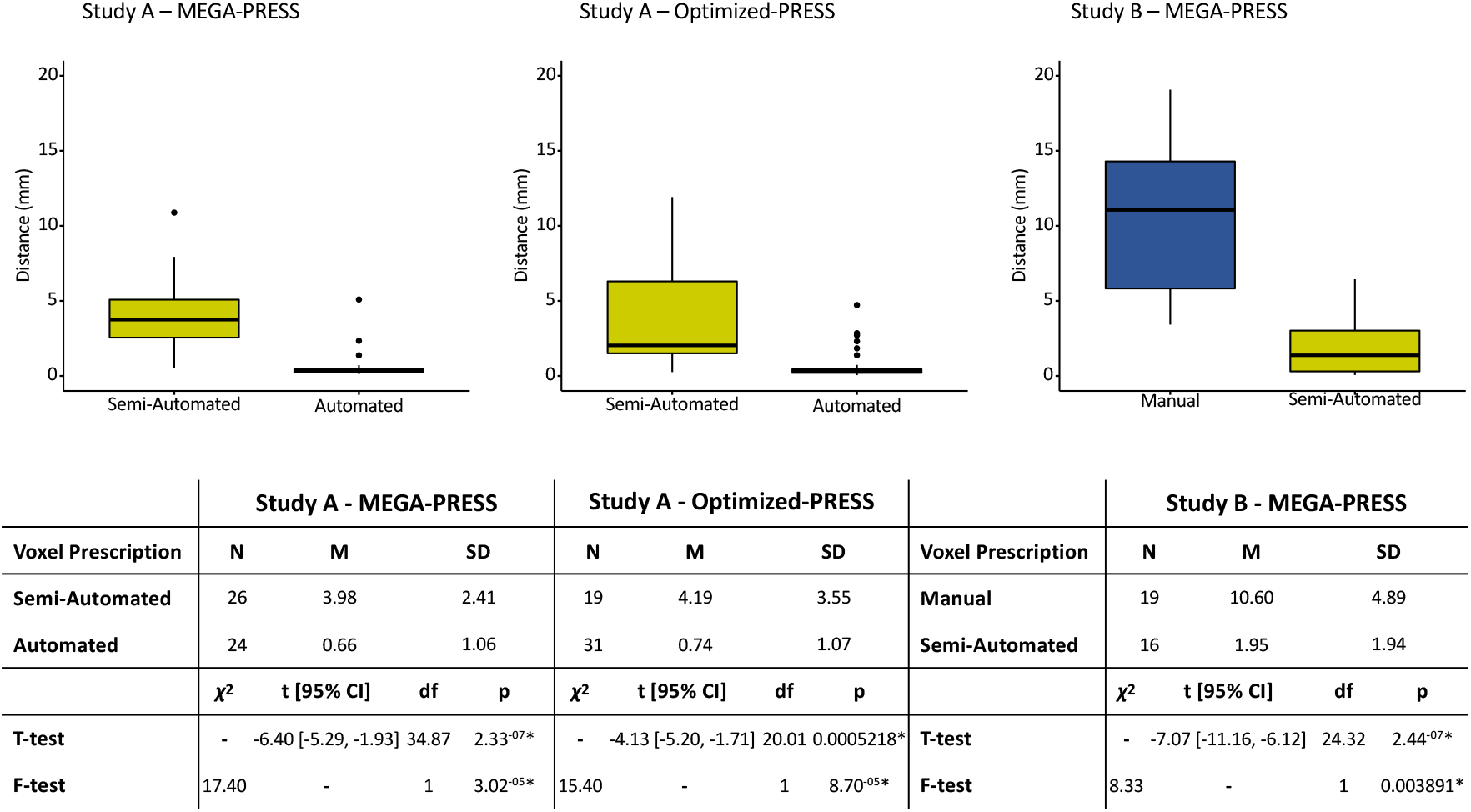
L-DLPFC Multi-Acquisition / Longitudinal Euclidean Distance: Automated voxel placement reduced variability and mean distance of center-of-gravity measurements. To determine the amount of change in geometric centers of prescriptions following repeated acquisitions, center-of-gravity coordinates were extracted for each voxel and Euclidean distance was calculated. Automated approaches reduced distance measurements across both MRS acquisition size and study cohorts.

## DISCUSSION

Here we report on novel voxel prescription pipelines that enable increasingly robust voxel prescription for multi-acquisition as well as longitudinal MRS applications. To evaluate the consistency of these approaches across several iterations of varying automation, we investigated the spatial and anatomical reproducibility of repeated MRS acquisition across two independent datasets. Validation of the technique was performed quantitatively by examining the change in Euclidean distance of the voxel center-of-gravity, defining the overlap of voxels using the DSC, and by evaluating the voxel tissue composition fraction across acquisitions. Results of both datasets are consistent, demonstrating less variance across repeated prescriptions with increasing automation of the prescription approach - even in data acquired over a broad timeframe (i.e., one month between MRS acquisitions).

As personalized clinical interventions continue to gain traction, parallel improvements in methods used to measure clinical changes are necessary to identify underlying pathology and/or evaluate treatment course. Ultimately, the choice of acquisition method may depend on the research question at hand and particularly whether effects are expected to be focal or widespread. For example, it could be reasonably hypothesized that systemic pharmacological interventions exert brain-wide effects and thus automation of voxel prescription may not be warranted. Alternatively, the use of imaging acquisitions or modalities (i.e., functional, structural, positron emission tomography) to guide voxel placement would not only be useful for placement but also to provide biological or physiological justification for positioning within a structure. On the other hand, for investigations utilizing targeted neuromodulation paradigms such as transcranial magnetic stimulation, focused ultrasound, or focal drug release methods, the use of automated techniques that are not solely based on anatomical landmarks is critical.

In both studies, automated and semi-automated center-of-gravity Euclidean distance and DSC demonstrated highly reproducible voxel placements. While consistent manual voxel prescription has been reported in brain regions with well-defined boundaries and/or structural features (36), it is worth noting that the DLPFC results demonstrated here were observed using relatively small VOIs in the absence of a specific set of anatomical landmarks. This further underscores the utility of our approach for functionally defined regions-of-interest. In developing the voxel placement procedure, two versions of the pipeline were created, referred to as semi-automated and automated. These approaches are similar with the addition of one adjustment step at the end. This adjustment step is critical for MRS collected in superficial brain structures such as the cortex or immediately adjacent to a sinus to ensure that voxels are completely bound to brain tissue. In deep brain structures this adjustment step would likely not be necessary. Although neither study directly compared all three approaches (manual, semi-automated, and automated) head-to-head, we demonstrated that increased automation of the voxel prescription process achieves highly spatially consistent voxels with marked reductions in within-subject tissue fraction variability. The distance and spatial overlap analyses provide complementary data that provide empirical evidence in support of this conclusion. Both the mean and variability of the distance measurements in the semi-automated pipeline did not increase across studies, even with increased time between scans (i.e., 1-month vs. 1-hour), and thus demonstrate the utility of this method across multi-acquisition and longitudinal study designs.

A source of variability not specifically accounted for in the outlined automation approaches is voxel rotation and is an area that warrants ongoing methodological development. To overcome this the present studies, the slope of the skull in the sagittal plane was used to guide voxel rotation, however, users will need to establish a set of criteria for consistent voxel rotation. The proposed voxel-placement methods do not completely obviate this challenge and source of variability from manual input particularly for subcortical VOIs. While the Euclidian distance comparison does not account for this potential source of variability as it is based on voxel center of gravity, the DSC does. This is because DSC determines the overlap of two volumes (here binary masks) that is thus useful for determining the effects that inconsistencies in rotation may have on the prescription pipeline. Our DSC results are consistent with a previously reported automated voxel prescription method that guides voxel placement based on anatomical ROIs (15), however, in the latter approach resampling is required. Resampling necessitates a modified calculation of the overlap coefficient that is termed the *generalized dice coefficient*. Additionally, Bai and colleagues assessed the reproducibility of manual voxel prescription by prescribing and acquiring the MRS voxel multiple times within a single scan session (i.e. patients were not removed from the scanner between acquisitions; (36)). Here we demonstrate that the spatial consistency of the automated and semi-automated approaches across multiple imaging sessions yielded greater inter-subject overlap coefficients comparatively even given the potential variability introduced by non-automated voxel rotation.

Consistency in data collection is critical for methodologies used in any research application and if not it begs to question the validity of the measurement. MRS is currently one of the only non-invasive imaging techniques that can measure neurochemical concentrations *in vivo*. Reducing unwanted sources of variability during MRS voxel prescription will lead to more consistent and meaningful results in human neuroscience, particularly when focal interventions are being evaluated. For example, as metabolic molecule concentrations vary across tissue type (30,31), tissue concentration within the MRS voxel influences the acquired measurements (37). For example glutamate and glutamine concentrations have been shown to be higher in concentration within GM compared to WM (38) highlighting the advantage of reduced GM and WM tissue fraction variability observed using automated voxel placement approaches. Collectively, these findings strengthen the utility of MRS applications to examine multi-session/longitudinal MRS data, in relation to basic and clinical research questions including effects of interventions. Future investigation is warranted to determine whether the automated approaches adapted for single-session MRS reduce MRS tissue fraction variability. Alternatively, voxel placement grounded on biological or physiological data rather than standardized anatomical guidance, may provide a more useful study measure, even if it increases between-subject spatial variability in voxel location.

Finally, reproducibility in neuroimaging is critical for both research and clinical applications. MRS is particularly susceptible to scrutiny on this front using standard voxel prescription methods that are dependent on user expertise. Automated pipelines, such as those described here, promise to broaden the applicability and generalizability of MRS. This is especially applicable for large-scale multi-center trials and investigations that are becoming increasingly common. In the current study, independent research staff without expertise in neuroanatomy acquired both independent datasets. The consistency of results among multiple users is strong evidence of the methods ease of use and demonstrates the applicability to standardize voxel placement across laboratories and institutions.

### Caveats

The current study evaluated the utility of using pre-determined functional ROIs generated from previous independent imaging sessions to guide the automated placement of MRS voxels. This requires an established analysis protocol that can be completed in a relatively short timeframe prior to follow-up imaging session. This may not be practical for all investigative teams. Real-time resting state analyses methods would enable the acquisition and analysis of functional imaging paradigms during the same session, and while frequency drift has been noted as a major concern for running gradient intensive sequences prior to collecting MRS, recent largescale data suggest that few scanners exhibit moderate to severe drift following fMRI using echo planar imaging (39). Alternatively, the automated approaches proposed in this manuscript could be easily adapted to perform multi-acquisition and longitudinal placement using coordinates from structural ROIs or following manual placement, although not formally tested in this manuscript. That is, manual prescription could be implemented initially, and the center-of-gravity coordinate of the MRS voxel could be documented and input into the co-registration steps for subsequent scans to achieve consistency of placement across repeated acquisitions.

As described above, a source of potential variability not fully accounted for with the proposed automated approaches are differences in voxel rotations across acquisitions. That is, centering the voxel prescription based on a center-of-gravity coordinates does not provide spatial information to ensure perfect overlap of the subsequent voxel prescription. Fortunately, discrepancies can be mitigated by establishing standard protocols for prescribing the rotation of the voxel. For example, cortical regions may be aligned to the slope of the skull in a designated anatomical plane. Future studies may develop increasingly automated procedures that algorithmically compute voxel rotation parameters based on anatomical properties (e.g., skull geometry).

### Summary and Conclusions

Our results provide evidence for the reliability and reproducibility of two pipelines that enable real-time automated MRS voxel prescription over multi-acquisition and longitudinal experimental approaches. The complimentary analyses and associated results highlight the utility of our approaches compared to manual procedures, that is: **(1)** greater consistency of tissue fraction within MRS voxels; **(2)** the reduction of distance between center-of-gravity measurements; and **(3)** substantial overlap as measured by the DSC across multiple users and projects. Together these results suggest that our approach provides a meaningful step toward the standardization of MRS data acquisition that is relevant for a variety of MRS research designs that consist of multiple users and laboratories. Our approach reduces the reliance on technician expertise during MRS data acquisition by standardizing voxel prescription and thus broadens the usability and feasibility of MRS as an investigative tool in neuroscience.

## Data Availability

The data and code that was used to generate the findings of this study is available upon request from either James Bishop, Ph.D. (Data) and Matthew Sacchet, Ph.D. (Code).

## ACKNOWLEDGEMENTS

We would like to thank Meng Gu, Ph.D., Ralph Hurd, Ph.D., and Adam Kerr, Ph.D. for their assistance and expertise with MRI data collection as well as Keith Sudheimer, Ph.D. for providing functional MRI analysis support in the ancillary studies. Declarations of interest: none. This work was supported by the NIH National Center for Complementary and Integrative Health grants 5R33AT009305-03 (NW & DS), 1F32AT010420-01 (JB), The Stanford University Molecular Imaging Scholars Fellowship (T32CA118681; JB), the Center for Neurobiological Imaging Innovation Award (JB), and the Phyllis and Jerome Lyle Rappaport Foundation (MDS). Additional data were provided by the Brain Stimulation Laboratory directed by (NW) and supported by Charles R. Schwab, the Gordie Brookstone Fund, the Marshall & Dee Ann Payne Fund, the Avy L. and Robert L. Miller Foundation, a Stanford Psychiatry Chairman’s Small Grant, NARSAD Young Investigator Award, and the Stanford Department of Psychiatry and Behavioral Sciences.

